# The Global case-fatality rate of COVID-19 has been declining disproportionately between top vaccinated countries and the rest of the world

**DOI:** 10.1101/2022.01.19.22269493

**Authors:** Najmul Haider, Mohammad Nayeem Hasan, Rumi A Khan, David McCoy, Francine Ntoumi, Osman Dar, Rashid Ansumana, Md. Jamal Uddin, Alimuddin Zumla, Richard A Kock

## Abstract

Globally 58.83% human population received at least one dose of the COVID-19 vaccines as of 5 January 2021. COVID-19 vaccination rollout is progressing at varied rates globally and data on the impact of mass vaccination on infection and case-fatality rates require definition. We compared the global reported cumulative case-fatality rate (rCFR) between top-20 countries with COVID-19 vaccination rates (>125 doses/100 people) and the rest of the world, before and after commencement of vaccination programmes.

We considered the 28th day of receiving the first vaccine in the world as a cut-off to compare the pre-vaccine period (Jan 1, 2020 – Jan 5, 2021) and the post-vaccine period (Jan 6, 2021-Jan 5, 2022). We used a Generalized linear mixed model (GLMM) with a beta distribution to investigate the association between the CFR and potential predictors of each country and reported the relative risk (RR) of each variable.

The mean rCFR of COVID-19 in the top-20 countries with vaccination rates was 1.83 (95% CI: 1.24-2.43) on 5 Jan 2021 and 1.18 (95% CI: 0.73-1.62) on 5 Jan 2022. The CFR for the rest of the world on 5 Jan 2021 was 2.32 (95% CI: 1.86-2.79) and 2.20 (95% CI: 1.86-2.55) on 5 January 2022. In Sub-Saharan Africa, the CFR remained roughly unchanged at 1.97 (95% CI: 1.59-2.35) on 5 Jan 2021 and 1.98 (95% CI:1.58-2.37) on 5 Jan 2022. The GLMM showed vaccination (/100 population) (RR:0.37) and Stringency Index (RR:0.88) were strong protective factors for the country’s COVID-19 CFR indicating that both vaccination and lockdown measures help in the reduction of COVID-19 CFR.

The rCFR of COVID-19 continues to decline, although at a disproportionate rate between top vaccinated countries and the rest of the world. Vaccine equity and faster roll-out across the world is critically important in reducing COVID-19 transmission and CFR.

**Key Questions:** 

**What is already known:**

- Vaccination can reduce the case-fatality rate of COVID-19. Globally, the COVID-19 vaccination rollout is progressing at varied rates.

**What are the new findings:**

- In the top-20 countries with vaccination, >200 doses of vaccines are given per 100 people on 5^th^ Jan 2022, In the rest of the word, the figure is 105, and in Sub-Saharan Africa (SSA) only 15.72
- After the introduction of COVID-19 vaccination the reported case-fatality rate (rCFR) of COVID-19 has reduced by 35% in the top-20 countries with vaccination, 8% in the rest of the world roughly unchanged in SSA.
- The doses of COVID-19 vaccines (/100 people) and rCFR has a negative correlation on 5 Jan 2022 (r=-0.296, p<0.001).
- The COVID-19 vaccination and Stringency Index are strong protective factors for the country’s COVID-19 rCFR indicating that both vaccination and lockdown measures help in reduction of COVID-19 rCFR.

**What do the new findings imply:**

- The disproportionate case-fatality rate of COVID-19 between top vaccinated countries and the rest of the world demand fast and equitable vaccine rollout globally to reduce COVID-19 transmission and CFR

## Introduction

In the early stages of the COVID-19 pandemic, the WHO-China Joint Mission on Coronavirus Diseases (COVID-19) 28 February 2020 report indicated a crude fatality ratio of 3.8% among the first 55,924 laboratory-confirmed cases ^1^. Subsequently, systematic reviews on the case-fatality rate (CFR) of COVID-19 reported an estimated CFR between 2.3-3.6% ^2–5^. The global cumulative reported case fatality rate (rCFR) of COVID-19 has increased up until the 17^th^ epidemiological week (April 22–28, 2020) of detection of SARS-CoV-2 in Wuhan China at 7.2, and then started to decline steadily up until 31 December 2021 at 2.2 ^6^. The decreasing rate of CFR has been explained by an increased rate of infection in the younger population or by the improvement of health care management, shielding from infection, and/or repurposing of several drugs that had shorted both hospitals stays and saved lives ^6^. However, during the last quarter of the year, 2020 different variants of concern/Interest (VOC/VOI) of SARS-CoV-2 started to emerge with increased transmissibility ^7^. Of them, Alpha (first detected in the UK in September 2020), Beta (first documented in South Africa in May 2020), Gamma (first detected in Brazil in November 2020), and Delta (first detected in India in October 2020) all reported having higher infectivity and severity than the virus originally detected in Wuhan, China ^7,8^. During November 2021, a new variant of concern, Omicron was reported to WHO (first reported from South Africa on 24 November 2021)^9^. Unlike previous VOC, Omicron shows a relatively lower case-fatality rate although the variant has a high transmissibility ^10,11^.

Vaccination can reduce the case-fatality rate of COVID-19. Several vaccines have been approved for emergency use by the Food and Drug Administration, USA, the European Medicine Agency, and the Public Health England and the World Health Organization approved use of a few Vaccines across the world. The real-world data had shown the effectiveness of the vaccines in terms of preventing infection by 60-92%^12,13^, hospitalization by 87-94%^12,14^ and deaths by 72%-100%^12,15^ after 7-28 days of receiving the second dose of the vaccines for Alpha, Beta, or Delta VOC.

Vaccines are not distributed equitably in the world. Although COVID-19 vaccines were developed at an unprecedented rate through the advancement of science and global cooperation, the distribution of the vaccine across the world is questionable ^16^. Current global vaccination rates of roughly 6.7 million doses per day translate to achieving herd immunity in approximately 4.6 years ^16^. Vaccine distribution is absent or very negligible in many of the low-income countries, and experts anticipated that 80% of the population in low-income countries would not receive a vaccine at the end of 2021 ^16^ which has been the case.

As the world enters the 3^rd^ year of the COVID-19 pandemic, globally, as of 13 January 2022, there have been over 315,3450, 967 confirmed cases of COVID-19, including 5,510,174 deaths, reported to WHO ^17^. COVID-19 vaccination rollout globally has progressed at varying rates and data on the impact of mass vaccination on infection and case-fatality rates require definition. To this end, we compared the global rCFR between the top-20 countries with vaccination rates (minimum vaccination: 125 dose/100 people) and the rest of the world, before and after commencement of vaccination programs. We further quantified the impacts of vaccinations, other control measures, and relevant variables on Covid-19 CFR.

## Methods

### COVID-19 data

The necessary COVID-19 related data, including daily new cases, daily new deaths, total deaths, and total deaths per million inhabitants, vaccination, were collected from the WHO daily COVID-19 situation reports of 210 countries from January 01, 2020, to January 05, 2022 ^18^. On 8^th^ December 2020, the first human in the world received an approved COVID-19 vaccine ^19^. We considered the 28th day of receiving the first vaccine in the world as a cut-off to compare the pre-vaccine period (Jan 1, 2020 – Jan 5, 2021) and the post-vaccine period (Jan 6, 2021-Jan 5, 2022).

### Reported case-fatality rate (rCFR)

We estimated daily cumulative rCFR COVID-19 (please read Hasan et al ^6^ for the definition of rCFR) as the number of deaths per 100 COVID-19 confirmed cases. In some instances, we aggregated the daily cases and deaths record by week. As the number of cases and deaths both are a fraction of total cases or deaths globally, we considered the term as reported CFR or to make a simplified version as rCFR ^6^ .

### Time series model to predict the trend

Three forecasting models (i.e., auto-regressive integrated moving average (ARIMA), automatic time-series forecasting model which is also known as ‘Prophet model’, and simple exponential smoothing (SES)), to identify the global trend of COVID-19 rCFR. We also used the Mann-Kendall (M-K) trend analysis to identify the presence of any trend and the direction of the trend (increasing or decreasing). We developed a generalized linear mixed model (GLMM) with beta distribution to identify whether the explanatory variables have any relationship between the country’s rCFR of COVID-19. These methods supported us to make a credible conclusion on the trend of COVID-19 rCFR over the time. All analyses were carried out using the statistical software R version 3.5.2.2 ^20^ and SAS ^21^.

We selected the SES, ARIMA, and Prophet Models because the key outcome variable (rCFR) is dependent on previous records (time-series events) and all these three models can take this into account. Using the time series models with the reported COVID-19 data, we forecasted trends for the prospective 10-days and visualized in the figure. SES is suggested as a good benchmark model to compare the performance of the ARIMA and Prophet models. We also used M-K trend analysis to identify the daily or weekly cumulative trend (increasing or decreasing) of COVID-19 rCFR ^22^. The details of the SES, ARIMA, and Prophet models are discussed in an earlier article on COVID-19 ^6^.

### Outcome and predictor variables

We collected a selected number of predictors variables from the World Bank and other UN sources such as population density ^23^, percentage of people above 65 years of age ^24^, Gross Domestic Product (GDP)^25^, worldwide governance indicators (WGI)^26^, and Global Health Security Index (GHSI)^27^, the prevalence of obesity ^28^ or from “Our World in Data” ^29^ in our analyses. We also included the country-specific prevalence of diabetes ^29^ and cardiovascular disease ^29^ to explain the variation of COVID-19 rCFR. The GHSI index scored between 0 and 100 to indicate the country’s capacity for early detection and reporting for epidemics ^27^. The WGI scored between -2.5 and 2.5, where -2.5 indicates the weakest and 2.5 indicates the strongest governance performance ^26^. The Oxford COVID-19 Government Response Tracker systematically collects information from 148 countries on several policy responses that governments have taken, scores stringency of such measures, and aggregates them into a common Stringency Index (SI) in a daily basis ^30^. In the Our World in Data ^29^, the SI was calculated by using nine response indicators including school closures, workplace closures, and travel bans, rescaled to a value from 0 to 100 (100 = strictest) ^30^. The value of the SI at any given day is the results of the mean of the nine indicators. Thus, the index reports a number that reflects the overall stringency of the government’s response. Higher index indicates a better overall response level ^30^.

### Empirical evaluation

We assessed the ARIMA and Prophet models by comparing their results to benchmarks model, the SES ^31^. The SES also allows the most appropriate non-seasonal model for each series, allowing for any kind of error or trend component. We, then, analyse and compared the performance of the performance of the time series models with some of the commonly used measures to evaluate the prediction significance including coefficient of determination (*R*^*2*^), root mean square error (RMSE), and mean absolute error (MAE).

### Generalized Linear Mixed Models

The Generalized Linear Mixed Models (GLMM) is an extension of the Generalized Linear Models (GLM) that allows the analysis of clustered categorical data, as in the case of repeated responses from different subjects ^32^. One of the key advantages of GLMM is that it separates the levels of the models to account for the group effect nesting the lower-level observations. In this study, locations are treated as the second level which group sequential observations within the same area, and independent variables are treated as repeated observations at the lower level. While the location data are assumed to be time-invariant, the independent data are assumed to be universal over the whole study area at a certain time point. The model describes a beta distribution family that has a logit link. We conducted GLMMs using the statistical software R ^33^.

### Statistical analysis

We performed summary statistical of vaccine doses/100 inhabitant and rCFR by country in the top-20 countries with vaccination rate, in SSA and in the rest of the world pre-and post-vaccination programme and reported the mean and inter-quantile range (IQR). We observed that the rCFR of COVID-19 has changed over time (**Fig. 1**). Using time-series models alone would not allow us to identify the reason behind the increasing and decreasing trend of COVID-19 rCFR. We explored whether the relationship between the rCFR of COVID-19 and country-level explanatory variables vary over time through generalized linear mixed models. As the trend of rCFR in both periods is different, we ran generalized linear mixed models to investigate the association between possible explanatory variables and tried to investigate which variables affect the most in both periods separately.

**Figure 1.**
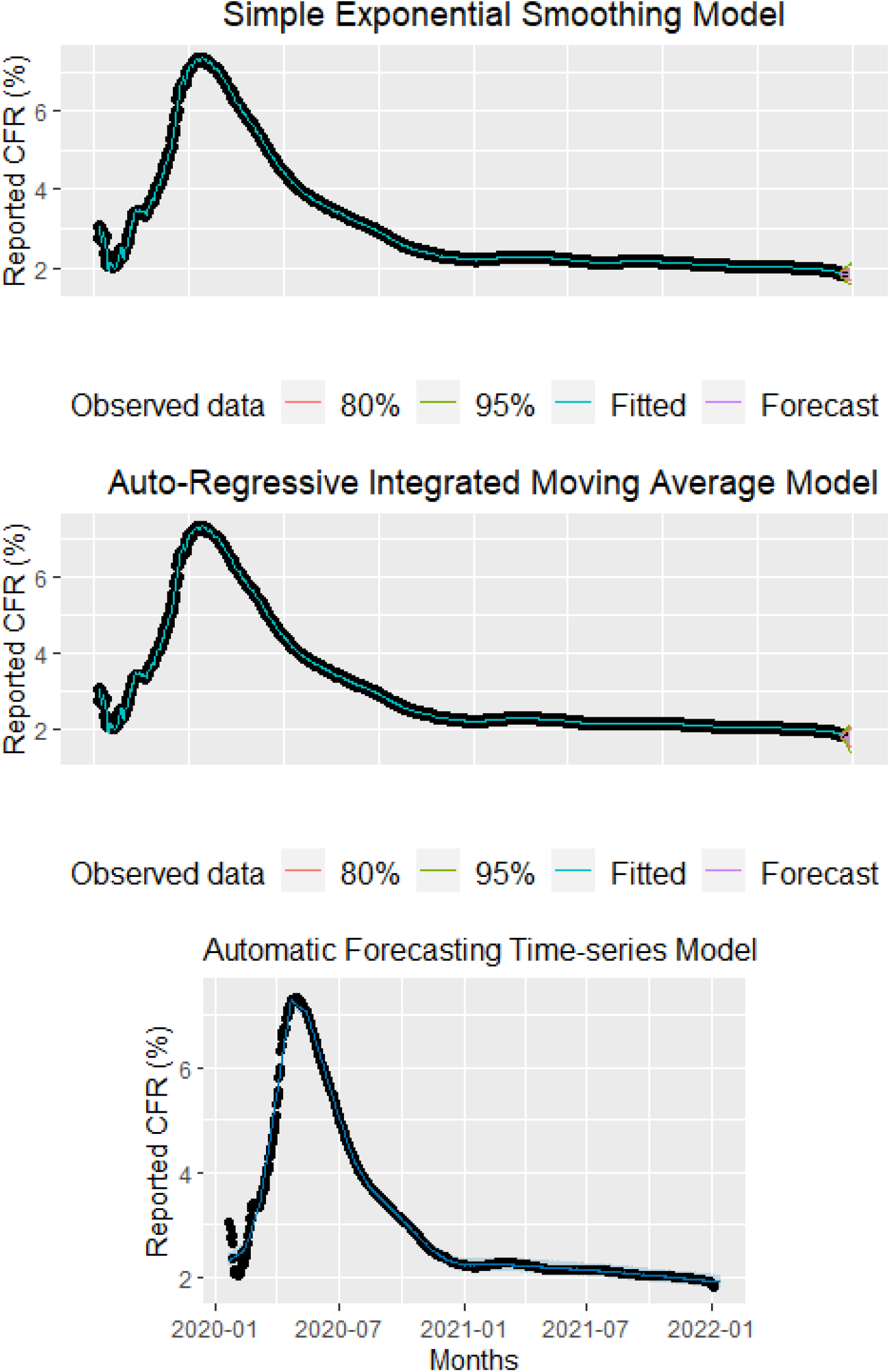
Top: Observed and predicted global daily reported Case-Fatality Rate (rCFR) using Simple Exponential Smoothing (SES) model. Middle: Observed and predicted daily worldwide daily cumulative rCFR using Auto-Regressive Integrated Moving Average (ARIMA) model. Bottom: Observed and predicted daily worldwide daily cumulative rCFR using Automatic Forecasting time-series model (Prophet model). The black dots indicate observed data, the blue line indicates the predictive CFR, and the shaded area indicates the 95% confidence interval of predicted CFR.

### Patient and Public Involvement

This study is based on different country’s COVID-19 cases and deaths record reported to WHO. The study does not include any individual patient level data. The findings of the results will be helpful in the equitable distribution of COVID-19 vaccines. The authors acknowledge the authorities shared the COVID-19 cases and deaths record. Patients or members of the public were not involved in the design, conduct, reporting or dissemination plans of the research as it was neither appropriate to involve nor possible to include.

## RESULTS

More than 297.77 million cumulative confirmed cases and 5.47 million deaths had been documented globally on 5 Jan 2022. More than 127.50 doses of COVID-19 vaccines are given per 100 population globally as of 5 January 2022. For every 100 people, 201.11 doses of vaccines are given in the top-20 countries with COVID-19 vaccination rates, in the rest of the world, 105.20 doses of COVID-19 vaccines are given, whereas the number is just 15.72 in Sub-Saharan Africa (SSA) **(Table 1)**. The global reported CFR is estimated as 1.83 on 5^th^ January 2021 which is has dropped by 35.5% at 1.18% on 5^th^ January 2022 in the top-vaccinated country. In the rest of the world the rCFR dropped from 2.32 on 5^th^ January 2021 to 2.2 on 5^th^ January 2022, reduced by only 8.4% only (**Table 1**). In the SSA, the rCFR has increased by 0.5% over the period (1.97 vs. 1.98). The top five countries with COVID-19 rCFR as of 5 Jan 2022 are Yemen (19.56%), Peru (8.75%), Mexico (7.44%), Sudan (7.07%), and Ecuador (6.09%). The correlating coefficient between vaccination rate (/100 people) and rCFR in different countries of the world on 5 Jan 2022 is estimated as -0.296 (p<0.001) **(Fig 2)**.

**Table 1:** The doses of COVID-19 vaccines and reported case-fatality rate (rCFR) of COVID-19 in the top-20 vaccinated countries and rest of the world during 5^th^ Jan 2021 and 5^th^ Jan 2022.

|  | Top-20 countries<br>with vaccination<br>rates<br>Mean (IQRs) | Rest of the<br>world<br>Mean (IQRs) | Global<br>Mean (IQRs) | Sub-Saharan<br>Africa<br>Mean (IQRs) |
| --- | --- | --- | --- | --- |
| Vaccination<br>Doses/100 People<br>(5 January 2022) | 201.11 (190.38 – 211.84) | 105.20 (91.28 – 119.11) | 127.50 (113.56 – 141.44) | 15.72 (35.31 – 26.12) |
| Reported CFR by 5 Jan 2021 | 1.83 (1.24 - 2.43) | 2.32 (1.86 - 2.79) | 2.26 (1.85 - 2.67) | 1.97 (1.59 – 2.35) |
| Reported CFR by 5 January 2022 | 1.18 (0.73 – 1.62) | 2.20 (1.86 – 2.55) | 2.07 (1.76 - 2.39) | 1.98 (1.58 – 2.37) |
| p-value (for differences of<br>rCFR between 5 Jan 2021<br>and 5 Jan 2022) | 0.001 | 0.340 | 0.069 | 0.972 |

**Figure 2:**
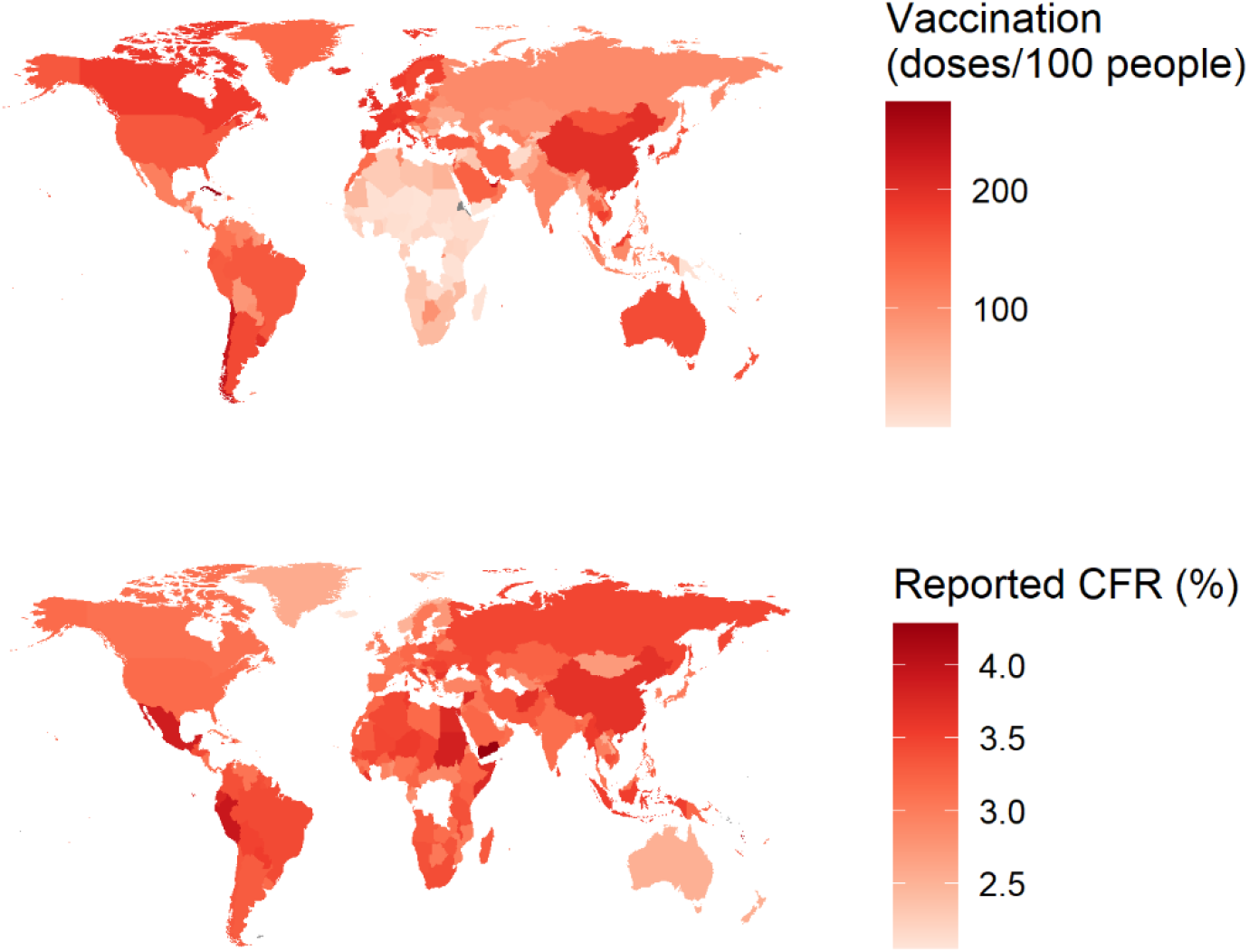
The map shows the Covid-19 vaccination rate (/100 people) and the reported Case-fatality rate (rCFR) in different countries of the world on 5 January 2022. The darker colour indicates the higher vaccination rates or higher case-fatality rates. An opposite correlation exists between COVID-19 vaccination rate and rCFR (r= -0.296 p<0.001)

### Factors associated with rCFR

In the GLMM, the estimated effect of each variable is presented in relative risk (RR) and its significance is shown by its p-value. The COVID-19 vaccination (0.37 [0.19–0.57]), GDP (0.75 [0.64–0.87]), and stringency index (0.88 [0.86–0.89]) was negatively significantly associated with the COVID-19 rCFR indicating that vaccination, GDP and lockdown measures all contributed to reduction of rCFR of COVID-19. The percentage of people aged 65 years or above the age of the population of the country were significantly positively (RR: 1.17, 95% CI: 1.01–1.37) associated with COVID-19 rCFR (**Table 2**).

**Table 2:** Factors associated with reported Case-Fatality Rate (rCFR) of COVID-19 in different counties of the world using a generalized linear mixed model during 1^st^ Jan 2020 and 5^th^ Jan 2022.

| Variables | Relative Risk [RR] | 95% Confidence Interval | P-value |
| --- | --- | --- | --- |
| Vaccination | 0.374 | 0.191 - 0.567 | <0.001 *** |
| The percentage of people aged 65 and above | 1.170 | 1.011 - 1.375 | 0.049* |
| Population density | 0.895 | 0.798 - 1.002 | 0.048* |
| GDP | 0.746 | 0.643 - 0.870 | <0.001 *** |
| GHSI | 1.123 | 0.955 - 1.321 | 0.150 |
| WGI | 1.050 | 0.906 - 1.213 | 0.500 |
| Obesity (%) | 1.121 | 0.993 - 1.269 | 0.060 |
| Stringency index | 0.882 | 0.864 - 0.887 | <0.001 *** |
| Vaccination: Weeks | 1.207 | 1.103 - 1.402 | <0.001 *** |
| Weeks | 0.914 | 0.817 - 0.955 | 0.015 ** |
| **Groups Name** | **Variance** | **Standard Deviation** |  |
| Location (Intercept) | 0.3354 | 0.5791 |  |
| Weeks (Intercept) | 0.0274 | 0.1655 |  |
| **Akaike information criterion (AIC)** | -201305.5 | **Conditional $R^2$** | 0.710 |
| **Bayesian Information Criterion (BIC)** | -201201.0 | **Marginal $R^2$** | 0.207 |
| **Root Mean Square Error (RMSE)** | 0.021 | **Intraclass correlation (ICC)** | 0.634 |

The table includes the various covariates and the random intercept in the model. The intraclass correlation coefficient (ICC) of 0.634 was calculated by dividing the variance of the random effect by the total variance. Thus, the spatial unit effects account for approximately 63.4% of the total variance of weekly rCFR, which suggests moderate reliability on location effects on weekly rCFR. It is also to be noted that with the introduction of a random intercept, “vaccination”, “population density”, “GDP”, “weeks” and “stringency index” had significant negative effects on weekly rCFR and “the percentage of people aged 65 and above” have significant positive effects.

### The Trend of global rCFR of COVID-19

In the ARIMA and Prophet Model, we found a strong declining trend of rCFR of COVID-19 between observed and predictive global rCFR of COVID-19 with an *R*^*2*^, RMSE, and MAE value of 99.94% and 99.56%, 0.04 and 0.13, and 0.01 and 0.06, respectively (**Table 3**). In terms of accuracy, the ARIMA model performed better over Prophet and SES model (with better *R*^*2*^, RMSE, and MAE value). The coefficient of determination of the ARIMA model was larger and errors are lower than Prophet and SES model. According to the forecast in both models, the ratio of COVID-19 rCFR is expected to decrease considerably in the coming 10 days (**Fig 1)**. In M-K trend analysis, we found a negative trend of cumulative rCFR (p <0.001 and tau = -0.79). In Sen’s slop test, the slope was -0.02 (95% CI: -0.03 to -0.01).

**Table 3.** The summary of Simple Exponential Smoothing (SES), Auto-Regressive Integrated Moving Average (ARIMA), Automatic forecasting time-series model (Prophet), Mann-Kendall (M-K), trend and Sen’s slope analysis. The SES, ARIMA, and Prophet models used daily cumulative reported case-fatality rate (rCFR) data whereas the M-K trend analysis and Sen’s slop used weekly cumulative rCFR data. The Kendall’s Tau value permits a comparison of the strength of correlation between two data series (here, week of the year 2020 and rCFR) ^22^.

| Method & Period | $R^2$ | RMSE | MAE |
| --- | --- | --- | --- |
| ***Simple Exponential Smoothing*** |  |  |  |
| Overall | 99.91% | 0.04 | 0.01 |
| ***Auto-Regressive Integrated Moving Average*** |  |  |  |
| Overall ARIMA (4,1,3) | 99.94% | 0.04 | 0.01 |
| ***Automatic Forecasting time-series model*** |  |  |  |
| Overall | 99.56% | 0.13 | 0.06 |
| ***Mann-Kendell trend analysis*** |  |  |  |
|  | **tau** | **P** |  |
|  | -0.79 | <0.001 |  |
| ***Sen's slop test*** |  |  |  |
|  | **Sen's Slope** | **95% CI** |  |
|  | -0.02 | -0.03 to -0.01 |  |

RMSE: Root Mean Square Error; MAE: Mean Absolute Error

## Discussion

The global rCFR of COVID-19 has been declining since May 2020 and the rate become plateaued or slightly increased with the emergence of different variants of concerns especially after the emergence and spreading of Delta VOC of SARS-CoV-2 ^34^. After the beginning of COVID-19 vaccines rollout, the rCFR started to decline, although, at a different rate in top-20 countries with vaccination rates, and the rest of the world. In SSA, where the vaccine rollout did not reach to a satisfactory mark yet, the rCFR of COVID-19 remained constant. Many factors affect the reduction of rCFR, however, our analysis indicates that the combination of vaccination, lockdown measures (stringency index), and the country’s GDP is contributing significantly to the reduction of rCFR. Earlier studies discussed potential role of an increased rate of infection in younger people or by the improvement of health care management, shielding from infection, and/or repurposing of several drugs had a beneficial effect on reducing the fatality rate of COVID-19. This analysis complements previous findings, however, generating immediate impact of inequitable vaccine rollout globally, indicating the importance of equitable distribution of COVID-19 vaccine to reduce fatalities due to COVID-19.

The negative correlation between doses of COVID-19 vaccines given and rCFR indicates the effectiveness of vaccines in reducing COVID-19 related deaths globally. The vaccines thus provide a pathway out of this pandemic, but strong, innovative policies that ensure fast and equitable distribution are absent ^16^. Vaccinating the world serves global interests of protecting each other’s health, and economy ^16^. Unless the vaccine reaches to a level of global herd immunity, these goals will not be accomplished ^16^. Our analysis showed a great difference of vaccine roll out in top vaccinated counties (201 doses/100 people) and the low-income countries (e.g. 15 doses/100 people in SSA) on 5 Jan 2022 ^35^. In high-income countries, third or even fourth dose of the COVID-19 vaccine programmes are ongoing ^36^ whereas more than 80% of people in the SSA did not receive a single dose of vaccine yet (as of 5 Jan 2022). The high-income countries organization (G7) nations have committed support for global vaccine procurement through the Covid-19 Vaccines Global Access (COVAX) program, which supplies vaccines to low-and-middle-income countries. Currently, COVAX planned to vaccinate at least 30% of the population of 92 low-and -middle income countries by the end of 2022 ^37^.

We found the COVID-19 vaccination, stringency index, and GDP as significantly associated with the reduction of rCFR of COVID-19. The major vaccines (mRNA, or adenovirus vectored) all are highly effective in reducing hospital admission and deaths ^35^, even though some of the vaccines were not very effective in limiting the infection ^35^. Thus, the vaccine rollout helped the high-income countries quickly reduce the burden of patients in hospitals thus limiting fatalities of COVID-19. However, in the rest of the world where vaccine rollout is still far from achieving herd immunity (70-80% of people receiving full doses of vaccines) the case-fatality rate has not declined significantly compared to that of the top vaccinated countries. However, in many countries especially in SSA, the natural infection had reached in a state to limit the infection and reduce the overall burden of the pandemic ^38^. In the Republic of Congo 66% of people in Brazzaville ^39^ in Malawi, 64.9% of blood donors ^40^ and in the Central African Republic, 74% of community residents had antibodies to SARS-CoV-2 ^41^.

Lockdown measures indicated as Stringency index has been associated with reduction of CFR of COVID-19. The Stringency index is a composite measure based on indicators including school closure, workplace closure, travel bans, and use of mask, and other social distancing practices. This has been associated with shielding vulnerable people from infection and contributes to the reduction of COVID-19 related fatalities. While lockdown has been shown beneficial in reducing cases and fatalities of COVID-19 in high-income countries ^42^, in Africa, lockdown has shown low or no benefit in terms of limiting transmissions ^43^. The country’s GDP is another indicator associated with the reduction of COVID-19 rCFR. This is believed that countries with higher national income deployed vaccines at a faster rate which reduce the local transmission and reduced rate of hospitalization thus allowing them to concentrate on the vulnerable population, all synergistically helping them reduce the fatalities of COVID-19. Earlier studies have also identified these variables as a risk factors/protective factors for mortality/fatality rate of COVID-19 ^6,44,45^.

Equitable distribution of SARS-CoV-2 vaccines is crucial to ending the COVID-19 pandemic ^46^. The circulation of the SARS-CoV-2 across the world among the large unvaccinated populations might allow the virus to reassort to become a new VOC. Furthermore, many animal species are susceptible to SARS-CoV-2 including Mink, Primates, rodents, cats, and dogs ^47^. Earlier studies showed the link of animal species in the generation of VOC including dogs for Alpha variant ^48^ and rodents for Omicron variant ^49^. Thus, this is important to reduce the circulation of SARS-CoV-2 in the human and animal populations to avoid further epidemics caused by new VOC. Equitable and faster vaccine rollout is the key to reducing the circulation of SARS-CoV-2 across the world.

## Limitation

The case, death record, and the number of people being tested are reported based on the number shared by countries of the world. However, different countries have different capabilities of testing populations, and countries used different strategies to test their population which has affected the reported cases and deaths figures. This ultimately affected our estimated CFR of COVID-19. However, this is a global issue, and we accept this limitation and thus refer to the CFR as reported CFR, not true Case-fatality rate. Also, this terminology should not be confused with the infection fatality rate. Although we found that vaccination and lockdown measures both are associated with the reduction of COVID-19 CFR, however, we could not differentiate the true impact of each measure separately although the RR of the vaccination is estimated as four times more effective compared to the lockdown measures.

## Conclusion

More than 201 doses of COVID-19 vaccines have been given per 100 people in the top-20 countries with higher vaccination rates, compared to 105 doses in the rest of the world and 16 doses in Sub-Saharan Africa as of 5^th^ Jan 2022. Vaccination and reported CFR (rCFR) of COVID-19 has a negative correlation (−0.29) on 5 Jan 2022 indicating the effectiveness of the vaccines in reducing COVID-19 related deaths. Globally, the reported CFR (rCFR) of COVID-19 has been declining since the beginning of the vaccination programme. However, the rCFR has dropped significantly in the top-20 countries with vaccination rates (35%) compared to the rest of the world (8%), whereas it remained largely unchanged in Sub-Saharan Africa (1.97 on 5^th^ Jan 2021 and 1.98 on 5^th^ Jan 2022). The COVID-19 vaccination, lockdown measures, and country’s GDP were associated with reduction of rCFR of COVID-19. Vaccine equity and faster roll-out across the world is critically important in reducing COVID-19 transmission and CFR.

## Contribution

NH and RK originally planned the study, MNH collected the data, NH and MNH, MJU analyzed the data. NH prepared the first draft manuscript and MNH, RAK, DM, FN, OD, RA, MJU, AZ and RK reviewed the draft manuscripts. All authors approved the submission of the manuscript.

## Data Availability

All the data presented in this manuscript are publicly available in "Our World in Data", "WHO" or "United Nations" webpages. However, the corresponding author can be reached to specific queries on the data sources.

## Acknowledgements

NH, FN, OD, RA AZ, RK are part of PANDORA-ID-NET Consortium (EDCTP Reg/Grant RIA2016E-1609) funded by the European and Developing Countries Clinical Trials Partnership (EDCTP2) programme, which is supported under Horizon 2020, the European Union’s Framework Programme for Research and Innovation.

## Ethical approval

This study does not include any individual-level data and thus does not require any ethical approval.

